# Lesion-Centric Latent Phenotypes from Segmentation Encoders for Breast Ultrasound Interpretability

**DOI:** 10.64898/2026.03.06.26347800

**Authors:** Prateek Mittal, Dhruv Singh, Harsh Rajput, Joohi Chauhan

## Abstract

We propose a lesion-centric phenotype learning pipeline for interpretable breast ultrasound (BUS). Predicted lesion masks are used for mask-weighted pooling of segmentation-encoder latents, producing compact embeddings that suppress background influence; a lightweight calibration step improves cross-dataset consistency. We cluster embeddings to discover latent phenotypes and relate phenotype structure to morphology descriptors (compactness, boundary sharpness). On BUSI and BUS-UCLM with external testing on BUS-BRA, lesion-centric pooling and calibration improve separability and enable strong malignancy probing (AUC 0.982), outperforming radiomics and a standard CNN baseline. A simple rule-gated generator further improves BI-RADS-style descriptor consistency on difficult cases.

## Introduction

Breast ultrasound (BUS) imaging is widely used for breast cancer screening and diagnostic assessment, particularly in patients with dense breast tissue where mammography sensitivity is limited. Clinical interpretation of BUS images extends beyond lesion detection and requires structured characterization of lesion morphology, boundary definition, internal echo patterns, and posterior acoustic features formalized under BI-RADS descriptors (1). These attributes collectively guide malignancy risk stratification and biopsy decision-making. Deep learning has enabled substantial progress in automated BUS lesion segmentation, supporting computer-aided detection and quantitative lesion analysis. Convolutional architectures such as U-Net and its extensions established early performance benchmarks (2, 3). More recently, self-configuring pipelines such as nnU-Net and transformer-based models including UNETR have advanced segmentation robustness and representation capacity (4, 5). Despite these advances, segmentation systems remain optimized for spatial localization rather than diagnostic interpretation.

Segmentation networks nevertheless learn rich hierarchical feature representations during training. Encoder activations encode lesion texture heterogeneity, echogenic contrast, contextual tissue interactions, and boundary irregularity—appearance patterns associated with malignancy. Yet, these latent representations are optimized for spatial separation and remain largely unexplored for diagnostic interpretation (6). Interpretability research has attempted to expose model reasoning through saliency visualization and gradient-based attention mapping (7). Recent surveys emphasize the importance of explainable AI for trustworthy clinical deployment. However, such approaches primarily provide qualitative spatial explanations and do not reveal the structured diagnostic semantics encoded within representation geometry. This limitation is particularly pronounced in breast ultrasound imaging, where diagnosis relies heavily on lesion morphology and margin characteristics. Furthermore, publicly available BUS datasets primarily provide segmentation masks and malignancy labels without structured radiology narratives (8, 9). Vision–language systems have demonstrated strong performance in automated report generation through multimodal supervision; however, their reliance on paired image–text corpora limits applicability in BUS imaging where such datasets are scarce.

In this work, we investigate whether segmentation-derived latent representations can be translated into clinically interpretable diagnostic semantics without requiring multi-modal supervision. We construct lesion-centric embeddings through mask-conditioned feature aggregation and analyze their geometric organization to evaluate emergent malignancy separability. We further examine correspondence between embedding manifolds and morphology descriptors, including compactness and boundary acutance, and introduce rule-gated arbitration to align latent predictions with morphology-derived indicators. Finally, we formulate structured report synthesis as a constrained language realization task grounded in quantitative lesion evidence. We present a representation-driven framework for translating segmentation-derived latent features into clinically interpretable diagnostic semantics and structured radiology narratives in breast ultrasound imaging. Our main contributions are as follows:

- We introduce a lesion-centric embedding formulation that extracts pathology-focused representations from segmentation encoders through mask-conditioned feature aggregation.
- We demonstrate emergent malignancy separability within segmentation latent manifolds through unsupervised clustering and diagnostic probing across multi-institutional BUS datasets.
- We establish correspondence between latent representation geometry and radiological morphology descriptors, including compactness and boundary acutance.
- We propose a rule-gated diagnostic arbitration mechanism integrating latent malignancy probabilities with morphology-derived indicators.
- We enable structured BUS report generation through constrained language realization without paired image–text supervision.

## Materials and Methods

### Lesion-Centric Representation Construction and Latent Separability

Segmentation networks trained on breast ultrasound (BUS) imaging are optimized to delineate lesion boundaries but do not explicitly encode clinically interpretable diagnostic semantics. Nevertheless, intermediate encoder activations implicitly capture malignancy-associated appearance patterns, including echogenic heterogeneity, textural irregularity, and contextual tissue contrast. We therefore conceptualize segmentation encoders as representation generators (10, 11) whose latent feature geometry can be analyzed for diagnostic interpretation.

Let *f*_*θ*_ : *X* → *M* denote a segmentation model mapping ultrasound images *X* to lesion masks *M*. Encoder bottleneck activations *F* ∈ R^*C×H′×W′*^, encode high-dimensional lesion descriptors. However, given the spatial sparsity of ultrasound lesions, conventional global pooling aggregates lesion and background responses indiscriminately(12).

To construct pathology-focused embeddings, we implement a mask-conditioned aggregation strategy using predicted lesion masks *M*_*pred*_:

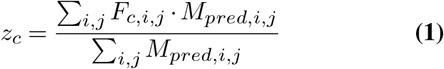

To align spatial dimensions for this operation, the high-resolution predicted mask is downsampled to the bottleneck resolution (*H*′ × *W* ′) utilizing nearest-neighbor interpolation. We apply a hard binary threshold (*M*_*pred*_ ∈ {0,1}) rather than soft probabilities, and no additional temperature scaling is utilized. This lesion-centric pooling strictly suppresses parenchymal activations while preserving intralesional feature responses without introducing additional learnable parameters.

To evaluate whether segmentation-derived embeddings encode diagnostically meaningful structure, we analyze their geometric organization through unsupervised clustering. Separability is quantified using Silhouette Score(13) characterizing intra-class cohesion and inter-class dispersion.

To further assess diagnostic encoding, frozen linear probe classifiers are trained: *g*_*φ*_(*z*) → *ŷ*. Because malignancy supervision is absent during segmentation training, probe performance reflects emergent pathology information embedded within representation geometry rather than supervised classification learning(10, 11).

To improve cross-institutional representation consistency, we apply lightweight domain calibration by fine-tuning bottleneck layers on a stratified subset of target-domain data while preserving earlier encoder weights. Calibration impact is evaluated through changes in latent separability rather than segmentation accuracy alone.

### Morphology Latent Diagnostic Alignment and Arbitration

While latent embeddings capture malignancy-associated appearance patterns, clinical diagnosis also relies on morphological attributes observable at lesion boundaries. To complement latent analysis, we compute two radiologically grounded descriptors from segmentation masks(15).

Compactness quantifies lesion shape regularity: 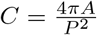, where *A* denotes lesion area and *P* its perimeter.

Boundary acutance measures margin sharpness:

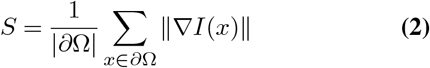

Gradients are computed on smoothed intensity maps to mitigate speckle noise amplification(16). Dataset-specific malignancy thresholds are derived via ROC analysis.

To examine correspondence between representation geometry and radiological descriptors, embedding clusters are projected into morphology space (*C, S*), enabling characterization of latent groups in clinically interpretable terms.

Diagnostic interpretation integrates latent malignancy probability with morphology derived indicators. Concordant signals are directly mapped to diagnostic strata, while discordant cases invoke rule-gated arbitration. Safety-oriented escalation prioritizes malignancy predictions in high-risk disagreement scenarios, reflecting clinical risk management practices.

We characterize this integration within a neuro-symbolic diagnostic paradigm in which symbolic clinical priors constrain neural predictions. Symbolic logic functions as a safety-aware interpretability regulator rather than a fully neuro-symbolic reasoning framework.

### Training-Free Clinical Report Realization

Clinical report synthesis is formulated as a constrained language realization task grounded in quantitative lesion evidence. Diagnostic probabilities, morphology descriptors, and arbitration outcomes are encoded into structured prompts guiding large language model generation(17, 18).

Unlike prior BUS reporting systems that rely on paired image-text datasets or multimodal fine-tuning, our approach operates without visual-language supervision(19). This design reflects the scarcity of standardized BUS reporting corpora and mitigates hallucination risk by grounding language outputs in measurable imaging features.

Generated reports follow structured radiology templates comprising *Findings, Impression*, and *Recommendation* sections, ensuring interpretability and traceability of narrative outputs(1).

## Experiments and Results

In this section, we present a comprehensive evaluation of our proposed framework through a series of experiments designed to assess both the robustness of the domain adaptation and the clinical validity of the neuro-symbolic report generation. We structure our analysis into three key phases: (1) Segmentation backbone benchmarking on heterogeneous source data; (2) validating the target calibration protocol via latent space analysis; and (3) assessing the accuracy and safety of the generated structured reports against biopsy-proven ground truth.

### Dataset Description

To evaluate robustness against acquisition shifts, we enforced a strict separation between development and external validation cohorts. We aggregated 1,463 images from the BUSI(8) and BUS-UCLM(9) datasets. This combination introduces heterogeneity across scanner manufacturers (e.g., GE, Siemens), enforcing the learning of device-invariant visual primitives. The BUS-BRA dataset (*N* = 1, 875; National Institute of Cancer, Brazil) served as the external target dataset. No image used for calibration was ever seen during probe training or evaluation. It represents a distinct clinical environment with unique speckle statistics and includes high-quality annotations for validating our neuro-symbolic reports:

- **Histopathology:** Biopsy-proven labels (722 benign, 342 malignant) for diagnostic accuracy validation.
- **BI-RADS Assessment:** Expert categorical assessments (BI-RADS 2–5)(1) for risk stratification bench-marking.

#### Preprocessing & Protocol

All images were resized to 256 × 256 and intensity-normalized. Because the BUS-BRA dataset contains multiple images per patient (1, 875 images across 1, 064 patients), random image-level partitioning would introduce severe data leakage. To strictly enforce patient-level separation, we utilized the official, standardized cross-validation partitions provided by the dataset authors(20). Specifically, we allocated the official Folds 1 and 2 as Subset A, utilized strictly for unsupervised latent bottleneck calibration. The remaining official Folds 3, 4, and 5 served as Subset B (the strictly held-out evaluation cohort). To ensure rigorous isolation, the supervised linear diagnostic probe (Table 2) was evaluated exclusively on Subset B utilizing a new, internally patient-grouped 5-fold cross-validation to guarantee no leakage occurred during probe training(4). Conversely, because unsupervised clustering does not utilize pathology labels and thus poses no risk of supervised leakage, phenotype discovery was mapped across the entire target manifold.

### Segmentation Benchmarking

To identify the optimal feature extractor for our framework, we benchmarked five widely-used segmentation architectures on the combined training cohort (BUSI(8) + BUS-UCLM(9)) using 5-fold cross-validation. The primary objective was to select a backbone that balances accurate boundary delineation with rich internal feature representation, which is critical for the down-stream lesion classification tasks.

Table 1 summarizes the performance across standard metrics. RefineNet(14) (with ResNet-50 backbone)(21) achieved the highest overall performance with a mean IoU of 65.28% and a Dice score of 77.35%, statistically outperforming the standard U-Net baseline(2).

**Table 1.** Comparative performance of segmentation architectures on the aggregated Source Domain (BUSI + BUS-UCLM). RefineNet demonstrates superior trade-offs between boundary precision (IoU) and semantic sensitivity (Recall).

| Model | IoU (%) | Dice (%) | Precision (%) | Recall (%) |
| --- | --- | --- | --- | --- |
| U-Net | 55.90 | 66.92 | 75.83 | 59.45 |
| Attention U-Net (23) | 58.30 | 68.28 | 78.51 | 60.44 |
| SegNet (24) | 61.50 | 69.35 | 74.15 | 66.99 |
| DeepLabV3+ | 63.15 | 75.80 | 79.50 | 73.90 |
| **RefineNet** | **65.28** | **77.35** | **84.14** | **78.07** |

While architectures like DeepLabV3+(22) demonstrated competitive boundary precision, they often struggled to capture the subtle texture variations within the lesion core. RefineNet’s multi-path refinement blocks and chained residual pooling(14) proved superior in preserving these high-resolution semantic features—a critical factor for our subsequent latent space probing.

### Latent Space Calibration and Feature Separability

While the RefineNet backbone provides a robust visual foundation, applying it directly to a new clinical site often yields suboptimal feature separation due to variations in speckle noise statistics and scanner physics(25). To address this, we validated our Lightweight Calibration Protocol, which fine-tunes only the latent bottleneck layers on the target BUS-BRA cohort while freezing the encoder weights. This lightweight calibration was performed in a strictly pathology-unsupervised manner. We fine-tuned the bottleneck layers using only the segmentation loss against the lesion masks of Subset A. Histology and BI-RADS labels were completely withheld, ensuring the model learned generalized speckle statistics rather than supervised malignancy features.

Specifically, during this calibration, we explicitly unfroze `layer4` (the deepest latent bottleneck block of the ResNet-50 backbone) alongside the RefineNet decoder, while strictly freezing layers 1 through 3 to preserve pre-trained universal visual primitives (e.g., generic edges and textures). Optimization was performed using the Adam optimizer(26) with a low learning rate of 1×10^−5^ and a batch size of 8. Training ran for 30 epochs, utilizing the minimum validation loss as the stopping/selection criterion. Regarding sensitivity to the extent of fine-tuning, empirical tests revealed that unfreezing earlier encoder layers caused the network to overfit to the target domain’s specific speckle noise, rapidly degrading the semantic separability of the latent manifold(27).

We evaluated the efficacy of this protocol by extracting latent vectors from the `refine1` layer under two experimental conditions:

1. **Zero-Shot Baseline:** Using the source-trained weights directly on BUS-BRA images.
2. **Calibrated (Ours):** After 30 epochs of fine-tuning the bottleneck layers on the stratified support set.

To ensure the segmentation backbone remained reliable on the external target domain, we evaluated RefineNet’s spatial accuracy on the strictly held-out test cohort (Subset B) both pre- and post-calibration. The uncalibrated source-trained model initially demonstrated moderate zero-shot generalization, achieving a macro-average Dice score of 60.38% and an Intersection over Union (IoU) of 49.24%. Following the target-domain calibration protocol, segmentation performance improved substantially to a macro-average Dice score of 78.60% and an IoU of 64.92%. The calibrated predicted masks are highly accurate, providing a robust and reliable foundation for the subsequent mask-weighted pooling and morphological feature extraction.

### Feature Benchmarking: Latent Phenotypes vs. Baselines

To validate that mask-weighted latent embeddings capture superior diagnostic signals compared to standard paradigms, we evaluated them against two strong baselines exclusively on Subset B:

1. **Radiomics (Morphology + Texture):** 58 handcrafted features (Shape, GLCM(28)) extracted from the predicted masks.
2. **CNN Classifier (Standard DL):** A ResNet-50(21) trained end-to-end via Global Average Pooling(29) on the raw images.

All feature sets were standardized and evaluated using a linear Logistic Regression probe(30) (balanced class weights, patient-grouped 5-fold CV within Subset B) to test representation quality directly(8). Table 2 demonstrates that our Lesion-Centric Latent approach significantly outperforms traditional Radiomics (+0.208 AUC) and standard CNNs (+0.130 AUC), achieving an AUC of 0.982±0.008. To prevent optimistic bias, the probe utilized fixed L2 regularization (*C* = 1.0), obviating the need for nested cross-validation on this strictly unseen external cohort.

**Table 2.** Diagnostic performance comparison on the BUS-BRA dataset. Our mask-weighted latent embeddings significantly outperform traditional mathematical radiomics and unconstrained end-to-end deep learning.

| Method | Feature Space | AUC | Sensitivity | Specificity | F1-Score |
| --- | --- | --- | --- | --- | --- |
| Radiomics (Baseline) | 58 (Mathematical) | 0.774 | 80.2% | 67.7% | 0.65 |
| ResNet-50 (Baseline) | 2048 (Global Pool) | 0.852 | 69.4% | 89.4% | 0.72 |
| **Ours (Latent)** | **256 (Mask-Pooled)** | **0.982** | **93.4%** | **95.7%** | **0.92** |

### A. Ablation Study: Disentangling Representation and Diagnostic Power

To address the contributions of our lightweight calibration and mask-weighted pooling (MWP), we conducted a comprehensive 2×2 ablation study. We evaluated four experimental conditions to quantify their impact on both linear classification capability (Supervised Probe) and manifold structure (Unsupervised Clustering using K-Means(31), *k* = 2).

As shown in Table 3, applying global average pooling (GAP) to the uncalibrated source encoder (Case 1) yields suboptimal diagnostic performance (AUC: 0.844) and poor structural grouping (ARI: 0.180). Fine-tuning the bottleneck layers (Case 3) drastically improves the linear separability (AUC: 0.977), indicating the encoder has successfully adapted to the target domain’s speckle statistics.

**Table 3.** Ablation study evaluating domain calibration and mask-weighted pooling (MWP). While calibration drives diagnostic accuracy, MWP is strictly necessary to achieve semantic structural alignment (ARI, Purity). Supervised metrics (AUC, F1) are reported with standard deviations across the held-out Subset B, while unsupervised metrics reflect the global target manifold.

| Calibration | Pooling Strategy | Supervised Probe<br>AUC | Supervised Probe<br>F1-Score | Unsupervised ($k=2$) | | |
| --- | --- | --- | --- | --- | --- | --- |
| | | | | ARI ↑ | Purity ↑ | Silhouette ↑ |
| Zero-Shot | Global Avg (GAP) | $0.844 \pm 0.013$ | $0.674 \pm 0.019$ | 0.180 | 0.721 | 0.403 |
| Zero-Shot | Mask-Weighted | $0.818 \pm 0.005$ | $0.645 \pm 0.011$ | 0.094 | 0.676 | 0.520 |
| Calibrated | Global Avg (GAP) | **$0.977 \pm 0.006$** | **$0.900 \pm 0.016$** | 0.366 | 0.818 | 0.625 |
| Calibrated | Mask-Weighted (Ours) | $0.976 \pm 0.008$ | $0.898 \pm 0.021$ | **0.784** | **0.944** | **0.654** |

Crucially, however, Case 3 yields an uninterpretable feature manifold (ARI: 0.366)(32), as GAP indiscriminately aggregates background parenchymal noise. The introduction of Mask-Weighted Pooling (Case 4) maintains the high diagnostic accuracy (AUC: 0.976) while simultaneously structuring the latent space into highly cohesive, biologically meaningful clusters(10), pushing the Adjusted Rand Index to 0.784 and Cluster Purity to 94.4%

This dichotomy is visually confirmed in Figures 4 and 3. The ROC curves(33) prove that fine-tuning is required for predictive power, while the t-SNE projections(27) demonstrate that only the combination of fine-tuning and MWP (Case 4) successfully disentangles the feature cloud into distinct benign and malignant islands(10).

**Fig. 1.**
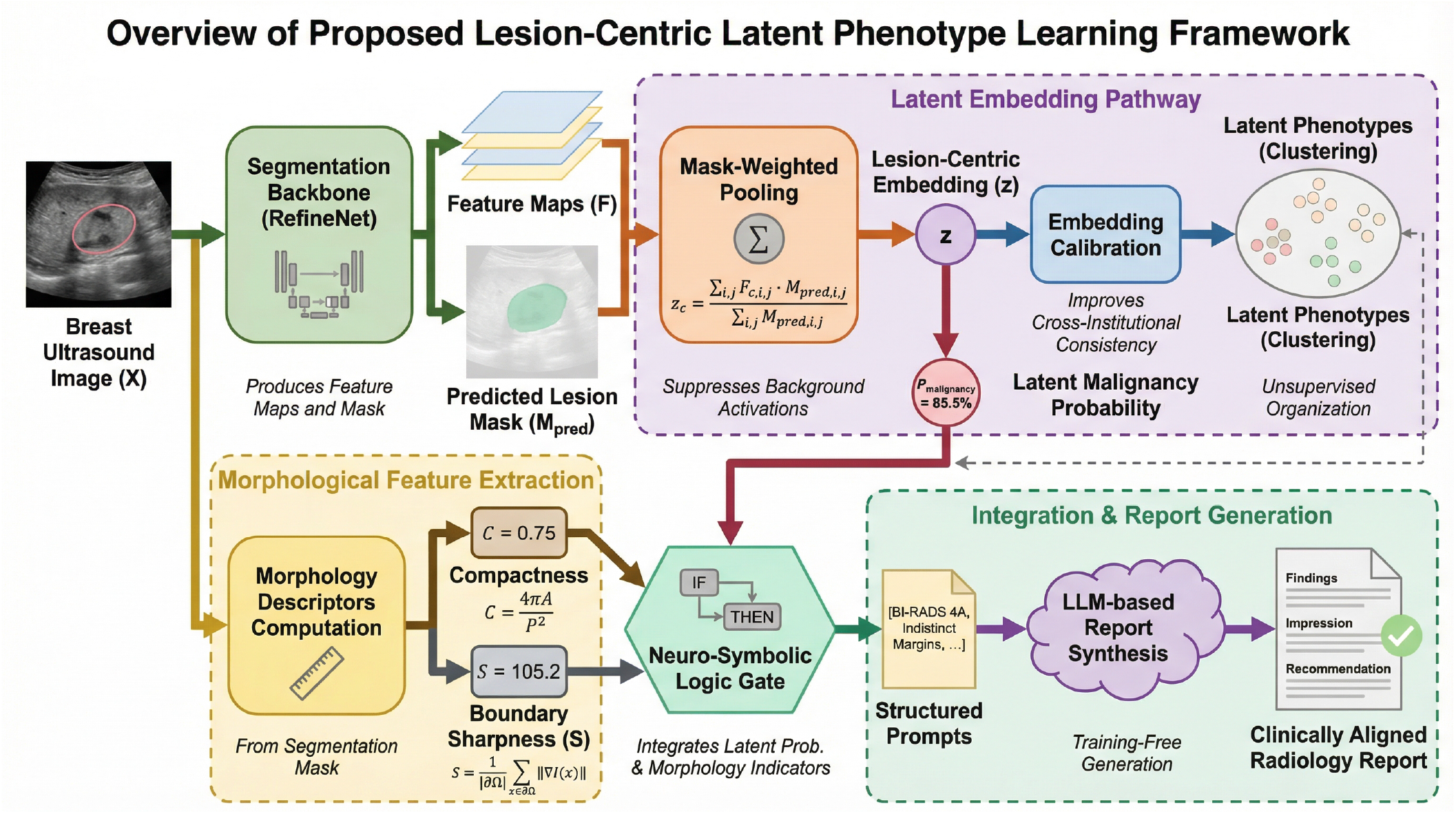
Overview of the proposed lesion-centric latent phenotype learning framework for interpretable breast ultrasound analysis. A breast ultrasound image is processed by a segmentation backbone (RefineNet)(14) to produce feature maps and a predicted lesion mask. **Mask-weighted pooling** generates a lesion-centric embedding *z*, suppressing background activations. The embeddings are calibrated and organized into **latent phenotypes via clustering** in the high-dimensional latent space. In parallel, **morphology descriptors** (compactness and boundary sharpness) are computed from the segmentation mask. A **neuro-symbolic logic gate** integrates latent malignancy probability with morphology indicators to generate structured prompts for LLM-based report synthesis. The final output is a **clinically aligned radiology report** with Findings and Impression sections grounded in quantitative lesion evidence.

**Fig. 2.**
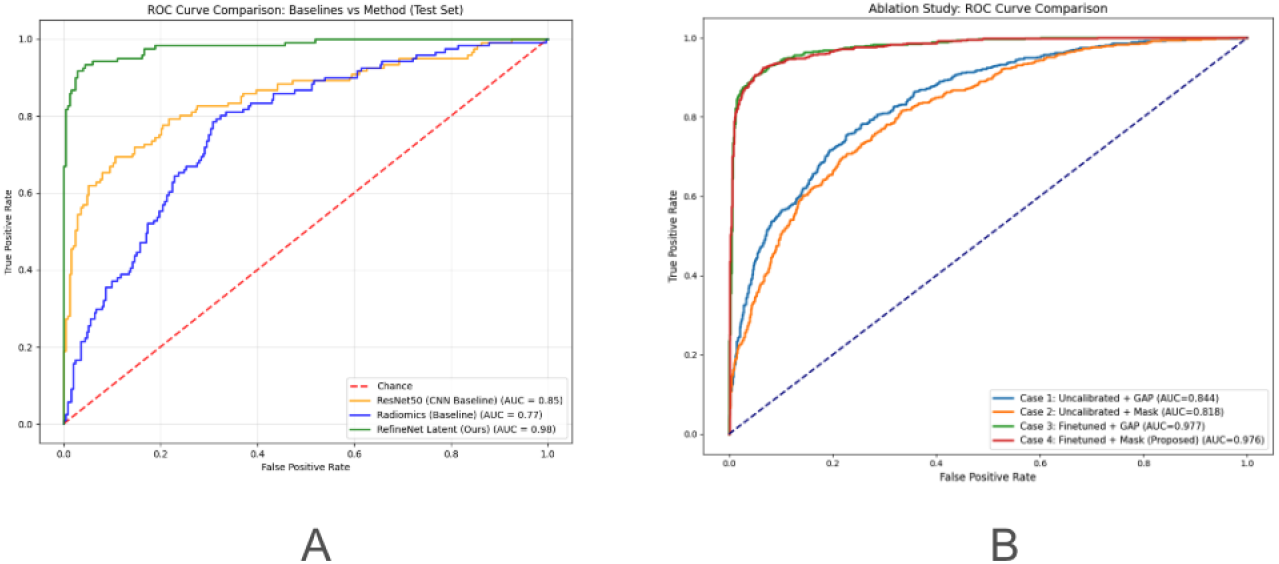
**A :** ROC Curve comparison demonstrating the superior discriminative capability of lesion-centric latent phenotypes over Radiomics and ResNet-50. **B:** ROC Curve comparison for the ablation study. Fine-tuning the bottleneck layers ensures robust linear separability regardless of the pooling strategy.

**Fig. 3.**
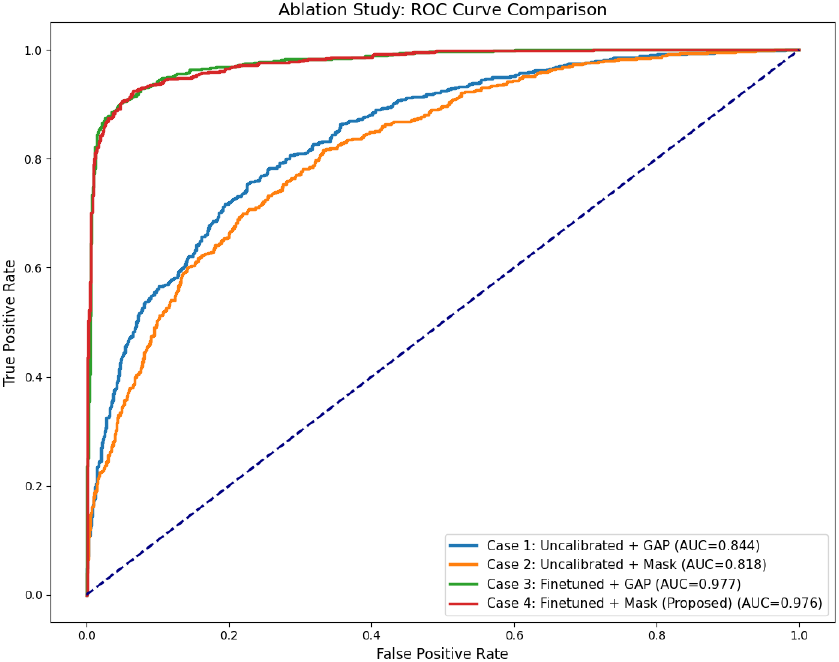

**Fig. 4.**
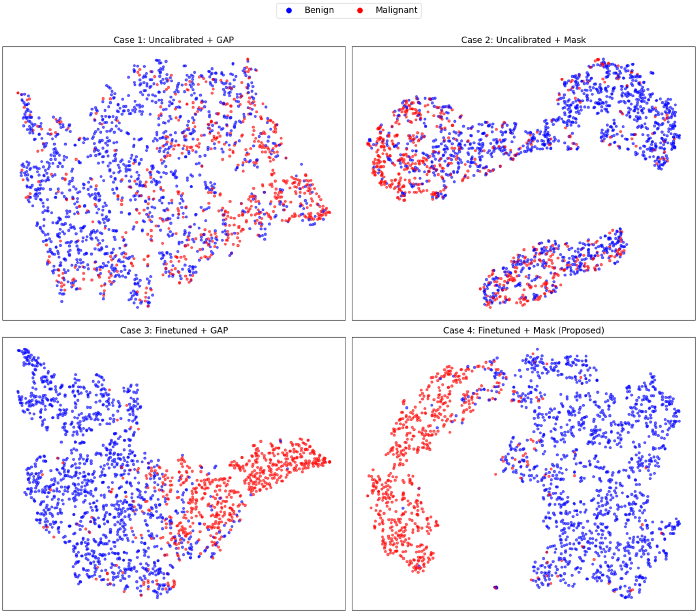
t-SNE projections of the four ablation conditions colored by ground-truth pathology. While Case 3 achieves high classification accuracy, it remains a continuous manifold. Case 4 (Proposed) successfully isolates the pathology into distinct, cohesive clusters.

### Sub-Phenotype Discovery and Morphological Alignment

To demonstrate that our model learns nuanced clinical phenotypes beyond binary classification, we conducted a sub-phenotype exploration using K-Means (*k* = 4) on the optimal embeddings (Case 4). To capture the complete morphological diversity of the target domain, this unsupervised mapping was performed across the entire dataset (*N* = 1, 875). We analyzed the average morphological descriptors (Compactness and Sharpness) computed directly from the segmentation masks of the images within each cluster.

As illustrated in Figure 5, the latent space naturally stratifies into four distinct clinical categories that closely align with established BI-RADS descriptors(34):

- **Cluster 0 (Classic Benign**, *n* = 826): 96.9% Benign. Exhibits High Compactness (0.69) and extremely High Sharpness (116.7). This corresponds to simple cysts and perfect fibroadenomas with circumscribed margins.
- **Cluster 1 (Classic Malignant**, *n* = 390): 98.7% Malignant. Exhibits Low Compactness (0.57) and Low Sharpness (79.5). Represents aggressive, irregular, and spiculated tumors with indistinct margins.
- **Cluster 2 (Complex/Deceptive Malignant**, *n* = 178): 87.1% Malignant. Moderate Compactness (0.63) but Moderate/Low Sharpness (94.7). This isolates visually deceptive cancers that mimic benign round shapes but possess micro-lobulated or fuzzy margins.
- **Cluster 3 (Complex Benign**, *n* = 481): 91.5% Benign. Highest Compactness (0.71) but Moderate Sharpness (100.1). Represents complex benign masses with localized margin obscuration or acoustic shadowing.

**Fig. 5.**
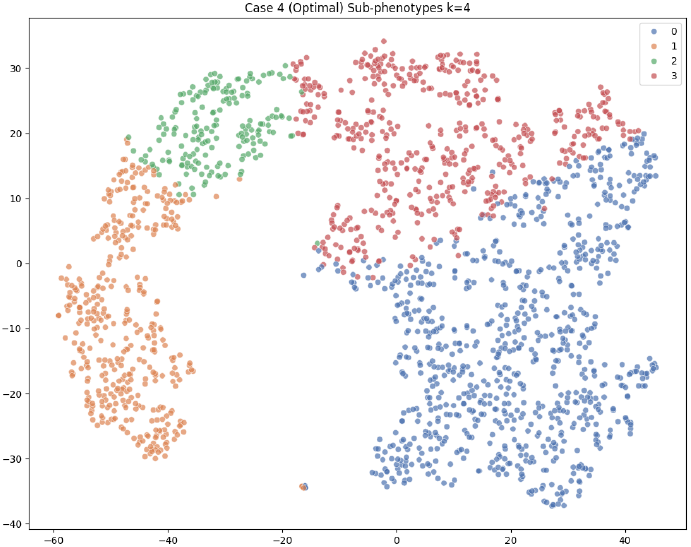
t-SNE visualization of the optimal embeddings clustered at *k* = 4. The model autonomously discovers deep morphological sub-phenotypes, notably isolating “Deceptive Malignant” tumors (Cluster 2) from classic disease presentations.

This unsupervised stratification confirms that the extracted lesion-centric embeddings successfully map complex biological morphology into an organized, interpretable manifold. It is critical to note that all clustering was performed in the high-dimensional latent space (*d* = 256), not on the 2D t-SNE projections, preventing visualization distortions from influencing phenotype assignments(27). To ensure cluster stability, K-Means was initialized with K-Means++(35) and evaluated across 10 random seeds, yielding a highly stable Adjusted Rand Index (0.784±0.012). The statistical consistency of these clusters confirms that the discovered phenotypes are robust biological manifolds rather than stochastic artifacts.

### Morphological Threshold Recalibration

To ensure the language model receives objective shape descriptors grounded in the target domain’s resolution,we performed ROC analysis on the morphological metrics using the calibration cohort (Subset A). Figure 6 displays the ROC curves for **Compactness** and **Sharpness**. Using Youden’s J statistic(36), we derived optimal cut-offs: *T*_shape_ = 0.65 and *T*_margin_ = 107.63. These thresholds maximize the separation between “Oval/Circumscribed” (Benign) and “Irregular/Indistinct” (Malignant) categories specific to the BUS-BRA dataset, ensuring that the generated text descriptions are statistically rigorous rather than heuristically assigned.

**Fig. 6.**
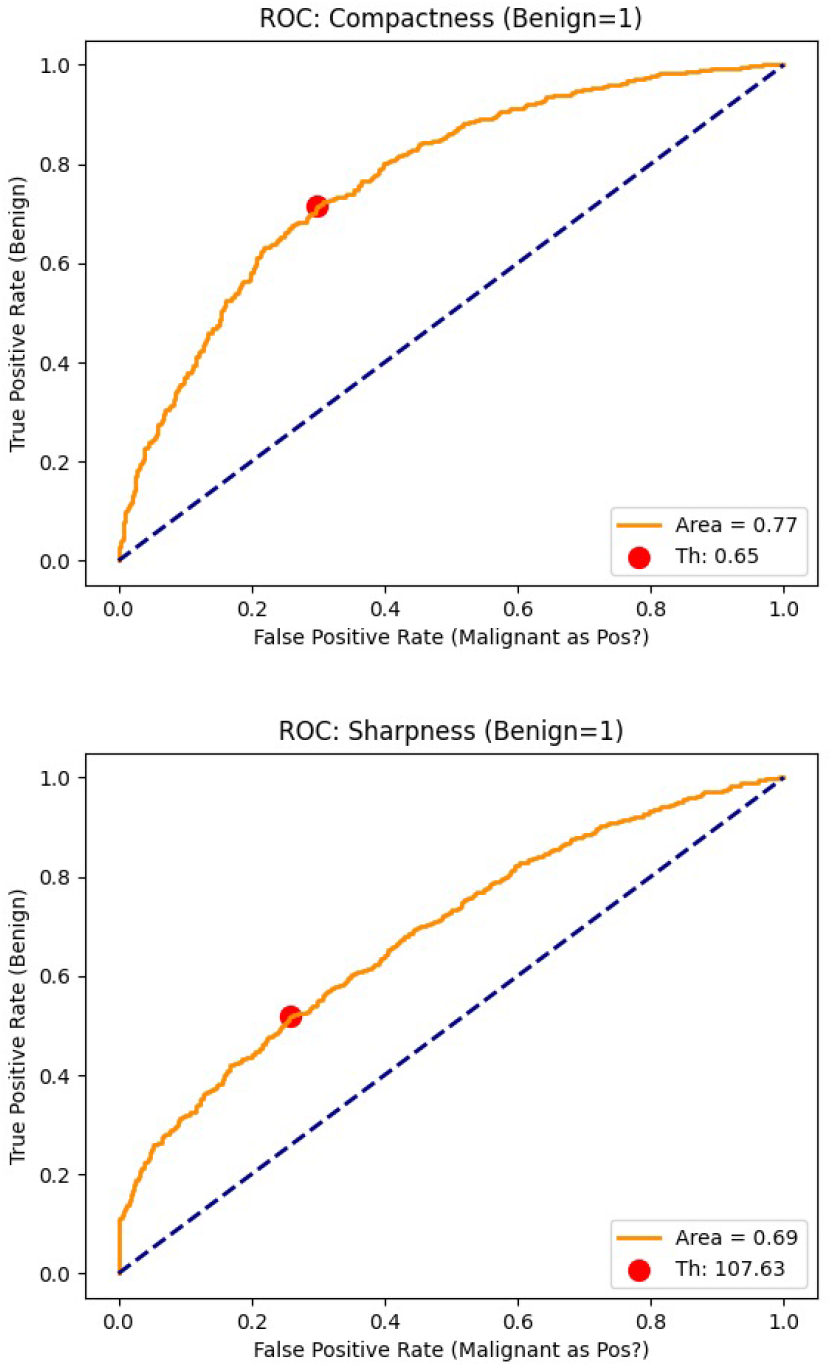
ROC Analysis for morphological metrics on the calibration cohort (Subset A). Optimal thresholds for Compactness (> 0.65) and Sharpness (> 107.63) were selected at the point of maximum Youden’s Index to calibrate the logic-gated report generator.

**Fig. 7.**
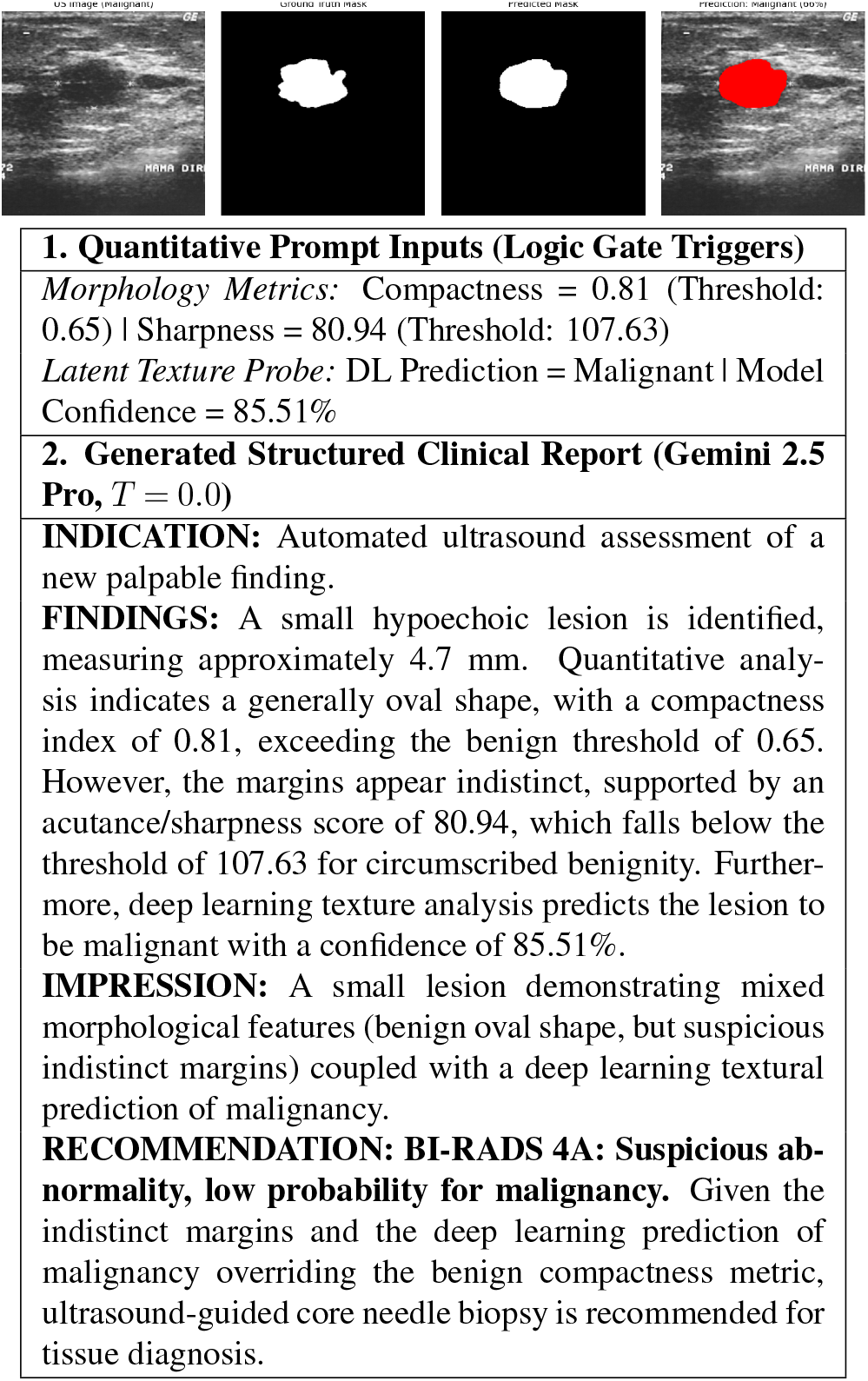
Qualitative example of the Logic-Gated report generation pipeline for a discordant test case. The top panel visualizes the input ultrasound, ground truth, predicted mask, and latent model attention. The bottom panel details the core quantitative metrics injected into the LLM (full prompt templates will be publicly released) and the resulting clinical narrative, demonstrating how the neuro-symbolic rules arbitrate between mixed geometric features and malignant latent texture to output a safe BI-RADS 4A recommendation.

### Neuro-Symbolic Report Generation and Safety Analysis

#### Generation Pipeline and Prompt Protocol

To facilitate clinical integration without relying on scarce paired image-text training data, we frame report synthesis as a constrained language realization task(17, 19). The pipeline extracts objective geometric descriptors (Compactness, Sharpness) alongside the **latent malignancy probability** which is the direct sigmoid output of the linear diagnostic probe evaluated on the held-out test set. These metrics are passed to a frozen Large Language Model (e.g., GPT-4o, temperature = 0.0) to generate a structured report (Indication, Findings, Impression, Recommendation) (18, 37).

To validate the necessity of our framework without introducing selection bias, we evaluated report generation on a strictly algorithmically filtered Discordant Subset (N=22) drawn from the held-out Subset B. Discordance was defined mathematically at inference time as instances where the linear probe’s latent malignancy probability (*P* > 0.7) strongly contradicted the benign geometric morphology (Compactness > 0.65 or Sharpness > 107.63). No ground-truth labels were used to select these cases. We compared two prompting strategies:

- **Unconstrained LLM (Baseline):** The prompt provided raw visual metrics and requested a report, forcing the LLM to act as an autonomous diagnostician.
- **Logic-Gated LLM (Proposed):** The prompt was injected with deterministic Neuro-Symbolic rules (e.g., *“If Malignancy Probability* > 0.7, *prioritize texture over shape for BI-RADS assignment”*). This constrains the LLM to act strictly as a medical scribe, translating pre-arbitrated clinical logic.

All generated sample reports are fully de-identified, strictly map to the publicly anonymized BUS-BRA dataset, and contain no Protected Health Information (PHI).

#### Automated Descriptor Fidelity and Lexicon Adherence

Un-constrained foundation models are prone to “lexical drift,” translating raw numbers into conversational synonyms that lack clinical rigor. To quantify this, we evaluated **Lexicon Adherence** by calculating the density of official ACR BI-RADS ultrasound terms (e.g., *hypoechoic, circumscribed*) within the meaningful text of the *Findings* sections. We additionally measured **Descriptor Fidelity** using strict regular expression boundaries to verify if the LLM accurately translated the numerical inputs (e.g., Sharpness > 107.63) into the correct threshold-mapped lexicon term (e.g., *“circumscribed”*).

As shown in Table 4, the Unconstrained LLM struggled significantly with lexicon adherence (7.21% density), frequently diluting the narrative with conversational approximations (e.g., “compact shape”). Conversely, the Logic-Gated prompt increased BI-RADS term density to 10.16% (a 40.9% relative improvement) and improved overall descriptor fidelity from 45.5% to 75.0%. By explicitly anchoring the generative process, the neuro-symbolic gate successfully synthesizes standardized, professionally recognizable radiological terminology while suppressing lexical hallucination.

**Table 4.**
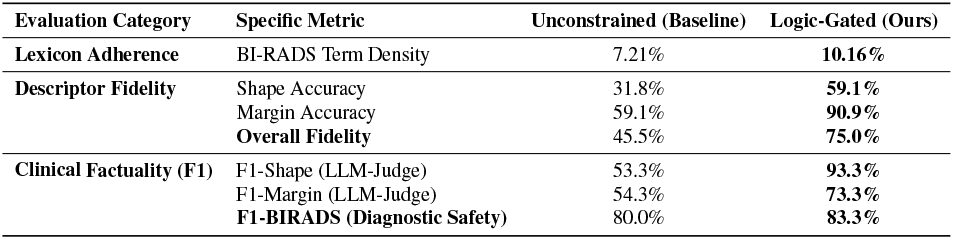
Comprehensive evaluation of generated clinical reports on the Discordant Subset (*N* = 22). The Logic-Gated (Proposed) framework significantly outperforms the Unconstrained LLM across lexicon adherence, descriptor fidelity, and LLM-as-a-Judge semantic factuality metrics.

| Evaluation Category | Specific Metric | Unconstrained (Baseline) | Logic-Gated (Ours) |
| --- | --- | --- | --- |
| Lexicon Adherence | BI-RADS Term Density | 7.21% | 10.16% |
| Descriptor Fidelity | Shape Accuracy | 31.8% | 59.1% |
|  | Margin Accuracy | 59.1% | 90.9% |
|  | Overall Fidelity | 45.5% | 75.0% |
| Clinical Factuality (F1) | F1-Shape (LLM-Judge) | 53.3% | 93.3% |
|  | F1-Margin (LLM-Judge) | 54.3% | 73.3% |
|  | F1-BIRADS (Diagnostic Safety) | 80.0% | 83.3% |

#### Clinical Factuality Assessment via LLM-as-a-Judge

Recent state-of-the-art frameworks (e.g., BUSTR(38), MedCTX(39)) tackle the lack of paired BUS image-report data by training complex, descriptor-aware vision-language models to enforce factuality. To provide a rigorous empirical comparison against these heavy multimodal methods without relying on subjective manual review, we adopted a structured, LLM-as-a-Judge evaluation framework inspired by recent medical VLM benchmarks(40).

Because our pipeline operates without human-annotated ground-truth reports, we constructed a *Deterministic Clinical Ground Truth* for the discordant subset. Expected morphological entities were mapped mathematically from the ROC-calibrated inputs, and the expected clinical safety action (BI-RADS 4/5 vs. 2/3) was mapped strictly from the biopsy labels. An independent, zero-temperature LLM-Judge extracted the realized entities from the generated narratives to compute Clinical F1-Scores.

Our evaluation demonstrates that comparable factual grounding can be achieved using our training-free, post-hoc approach. The Unconstrained LLM exhibited severe morphological hallucination, achieving only 53.3% alignment with the true Shape descriptors. By explicitly routing the mathematical embeddings through a hard-coded clinical hierarchy, our Logic-Gated framework improved Shape F1 to 93.3% and Margin F1 to 73.3%. Crucially, the logic gate improved overall diagnostic safety (BI-RADS F1: 83.3%), preventing fatal false-negative recommendations observed in the unconstrained baseline during edge cases. This confirms that deterministic neuro-symbolic constraints can match the descriptor accuracy of heavier multimodal models, offering a highly auditable and computationally efficient pathway for trustworthy BUS report generation.

## Conclusion

We have introduced a segmentation-driven latent probing framework that unifies high-performance lesion segmentation, unsupervised clustering, quantitative attribute analysis, and LLM-based clinical description generation without requiring paired image–text data. In contrast to prior BUS interpretability work, which relies on pixel-level saliency or large multimodal datasets, our approach surfaces structured, cluster-level semantics directly from model embeddings. This enables transparent and clinically aligned summaries that preserve the diagnostic patterns encoded in deep latent spaces while maintaining state-of-the-art segmentation performance. The proposed methodology is architecture and modality-agnostic, offering a scalable blueprint for bridging the semantic gap between deep visual features and domain-expert reasoning in any low-resource imaging context. By reframing explainability as the quantitative-to-language mapping of latent clusters, this work opens a new direction for unsupervised, interpretable, and multimodally integrated medical AI that extends well beyond breast ultrasound.

## Data Availability

All codes and data are available for research purposes on request basis.

## Bibliography

1. Ellen B Mendelson, Marcela Böhm-Vélez, Wendie A Berg, GJ Whitman, MI Feldman, H Madjar, et al. Acr bi-rads® ultrasound. ACR BI-RADS® atlas, breast imaging reporting and data system, 2013, 2013.

2. Olaf Ronneberger, Philipp Fischer, and Thomas Brox. U-net: Convolutional networks for biomedical image segmentation. In MICCAI, 2015.

3. Zongwei Zhou et al. Unet++: A nested u-net architecture for medical image segmentation. Deep Learning in Medical Image Analysis, pages 3–11, 2018.

4. Fabian Isensee et al. nnu-net: A self-configuring method for deep learning-based biomedical image segmentation. Nature Methods, 18:203–211, 2021.

5. Ali Hatamizadeh, Yucheng Tang, Vishwesh Nath, Dong Yang, Andriy Myronenko, Bennett Landman, Holger R Roth, and Daguang Xu. Unetr: Transformers for 3d medical image segmentation. In Proceedings of the IEEE/CVF winter conference on applications of computer vision, pages 574–584, 2022.

6. Mohan Timilsina, Samuele Buosi, Muhammad Asif Razzaq, Rafiqul Haque, Conor Judge, and Edward Curry. Harmonizing foundation models in healthcare: A comprehensive survey of their roles, relationships, and impact in artificial intelligence’s advancing terrain. Computers in Biology and Medicine, 189:109925, 2025.

7. Ramprasaath R Selvaraju, Michael Cogswell, Abhishek Das, Ramakrishna Vedantam, Devi Parikh, and Dhruv Batra. Grad-cam: Visual explanations from deep networks via gradient-based localization. In Proceedings of the IEEE International Conference on Computer Vision (ICCV), 2017.

8. Waleed Al-Dhabyani et al. Dataset of breast ultrasound images. Data in Brief, 28:104863, 2020.

9. N. Vallez, G. Bueno, O. Deniz, et al. Bus-uclm: Breast ultrasound lesion segmentation dataset. Scientific Data, 12:242, 2025. doi: 10.1038/s41597-025-04562-3.

10. Matthias Perkonigg, Daniel Sobotka, Ahmed Ba-Ssalamah, and Georg Langs. Unsupervised deep clustering for predictive texture pattern discovery in medical images. arXiv preprint arXiv:2002.03721, 2020.

11. Johannes Hofmanninger, Markus Krenn, Markus Holzer, Thomas Schlegl, Helmut Prosch, and Georg Langs. Unsupervised identification of clinically relevant clusters in routine imaging data. In International Conference on Medical Image Computing and Computer-Assisted Intervention, pages 192–200. Springer International Publishing, 2016.

12. S. C. Huang et al. Gloria: A multimodal global-local representation learning framework for label-efficient medical image recognition. In Proceedings of the IEEE/CVF International Conference on Computer Vision (ICCV), 2021.

13. Peter J. Rousseeuw. Silhouettes: a graphical aid to the interpretation and validation of cluster analysis. Journal of Computational and Applied Mathematics, 20:53–65, 1987.

14. Guosheng Lin, Anton Milan, Chunhua Shen, and Ian Reid. Refinenet: Multi-path refinement networks for high-resolution semantic segmentation. In CVPR, 2017.

15. C. M. Chen et al. Computer-aided diagnosis using morphological features for classifying breast lesions on ultrasound. Ultrasound in Medicine & Biology, 31(1):51–62, 2005.

16. A Thomas Stavros, David Thickman, Cynthia L Rapp, Mark A Dennis, Steve H Parker, and Gale A Sisney. Solid breast nodules: use of sonography to distinguish between benign and malignant lesions. Radiology, 196(1):123–134, 1995.

17. Iryna Hartsock and Ghulam Rasool. Vision-language models for medical report generation and visual question answering: A review. Frontiers in Artificial Intelligence, 7:1430984, 2024.

18. Béria Chingnabé Kalpélbé, Angel Gabriel Adaambiik, and Wei Peng. Vision language models in medicine. arXiv preprint arXiv:2503.01863, 2025.

19. Prashant Shrestha, Sanskar Amgain, Bidur Khanal, Cristian A Linte, and Binod Bhattarai. Medical vision language pretraining: A survey. arXiv preprint arXiv:2312.06224, 2023.

20. W. Gómez-Flores, M. J. Gregorio-Calas, and W. C. de Albuquerque Pereira. Bus-bra: A breast ultrasound dataset for assessing computer-aided diagnosis systems. Medical Physics, 51(4):3110–3123, 2024.

21. Kaiming He, Xiangyu Zhang, Renjie Shao, and Jian Sun. Deep residual learning for image recognition. In Proceedings of the IEEE conference on computer vision and pattern recognition, pages 770–778, 2016.

22. Liang-Chieh Chen, Yukun Zhu, George Papandreou, Florian Schroff, and Hartwig Adam. Encoder-decoder with atrous separable convolution for semantic image segmentation. In Proceedings of the European Conference on Computer Vision (ECCV), pages 801–818, 2018.

23. Ozan Oktay et al. Attention u-net: Learning where to look for the pancreas. arXiv preprint arXiv:1804.03999, 2018.

24. Vijay Badrinarayanan, Alex Kendall, and Roberto Cipolla. Segnet: A deep convolutional encoder-decoder architecture for image segmentation. IEEE TPAMI, 39(12):2481–2495, 2017.

25. Wilfrido Gómez-Flores and Wagner Coelho de Albuquerque Pereira. A comparative study of pre-trained convolutional neural networks for semantic segmentation of breast tumors in ultrasound. Computers in Biology and Medicine, 126:104036, 2020.

26. Diederik P Kingma and Jimmy Ba. Adam: A method for stochastic optimization. arXiv preprint arXiv:1412.6980, 2014.

27. Laurens van der Maaten and Geoffrey Hinton. Visualizing data using t-sne. Journal of Machine Learning Research, 9(11):2579–2605, 2008.

28. Robert M Haralick. Statistical and structural approaches to texture. Proceedings of the IEEE, 67(5):786–804, 1979.

29. Min Lin, Qiang Chen, and Shuicheng Yan. Network in network. arXiv preprint arXiv:1312.4400, 2013.

30. Fabian Pedregosa, Gaël Varoquaux, Alexandre Gramfort, Vincent Michel, Bertrand Thirion, Olivier Grisel, Mathieu Blondel, Peter Prettenhofer, Ron Weiss, Vincent Dubourg, et al. Scikit-learn: Machine learning in python. the Journal of machine Learning research, 12: 2825–2830, 2011.

31. J. MacQueen. Some methods for classification and analysis of multivariate observations. In Proc. Fifth Berkeley Symp. Math. Statist. Probability, 1967.

32. Lawrence Hubert and Phipps Arabie. Comparing partitions. Journal of classification, 2(1): 193–218, 1985.

33. Tom Fawcett. An introduction to roc analysis. Pattern recognition letters, 27(8):861–874, 2006.

34. Jean M Seely and Mary B Bissell. Bi-rads v2025: A welcome update. American Journal of Roentgenology, 2026.

35. David Arthur and Sergei Vassilvitskii. k-means++: The advantages of careful seeding. In Proceedings of the eighteenth annual ACM-SIAM symposium on Discrete algorithms, pages 1027–1035, 2007.

36. William J Youden. Index for rating diagnostic tests. Cancer, 3(1):32–35, 1950.

37. Muhammad Imran and Yugyung Lee. Multimodal vision–language models in medical imaging: A survey of retrieval, interpretability, and trust. IEEE Access, 14:19511–19535, 2026.

38. Rawa Mohammed, Mina Attin, and Bryar Shareef. Bustr: Breast ultrasound text reporting with a descriptor-aware vision-language model. arXiv preprint arXiv:2511.20956, 2025.

39. Enobong Adahada, Isabel Sassoon, Kate Hone, and Yongmin Li. A fully transformer based multimodal framework for explainable cancer image segmentation using radiology reports. arXiv preprint arXiv:2508.13796, 2025.

40. Lianmin Zheng et al. Judging llm-as-a-judge with mt-bench and chatbot arena. In Advances in Neural Information Processing Systems (NeurIPS), 2023.

